# The Negative Association between Dodecanoic Acid Intake and Cataract Incidence Based on NHANES 2005–2008

**DOI:** 10.1101/2025.03.20.25324346

**Authors:** Luo Jincheng, Xu Jingkai, Yang Yifeng, Li Doudou, Qiu Hongbin

## Abstract

**Objective:** This study aimed to investigate the relationship between different types of fatty acid intake and cataract incidence using data from NHANES 2005-2008, with the goal of providing scientific evidence for guiding fatty acid supplementation in the elderly and preventing cataracts.

**Methods:** A total of 2,536 participants were selected from the 2005-2008 National Health and Nutrition Examination Survey (NHANES) database. Student’s t-test and Pearson chi-square test were used to analyze the correlation between baseline data, characteristic data, and all dietary fatty acid intakes with cataract incidence. Weighted multivariate logistic regression was employed to examine the relationship between all fatty acid intakes and cataracts. Quartile regression analysis was used to evaluate the effect of dodecanoic acid on cataracts at different intake levels.

**Results:** After adjusting for covariates, significant differences were observed in the daily intakes of total fat (69.80 g *vs*. 62.46 g, p=0.005), total saturated fatty acids (23.06 g *vs*. 20.60 g, p=0.002), total monounsaturated fatty acids (25.53 g *vs*. 22.76 g, p=0.009), and total polyunsaturated fatty acids (15.16 g *vs*. 13.61 g, p=0.014) between the two groups. When cataract was used as the outcome variable, a significant negative correlation was found between daily dodecanoic acid intake and cataract in the weighted multivariate logistic regression model (Model 1: OR=0.79, 95% CI=0.65-0.97; Model 2: OR=0.80, 95% CI=0.65-0.97; Model 3: OR=0.81, 95%

CI=0.66-0.99). In the quartile regression analysis, the fourth quartile of daily dodecanoic acid intake was negatively correlated with cataract incidence in all models (Model 1: OR=0.58, 95% CI=0.39-0.86; Model 2: OR=0.58, 95% CI=0.39-0.86; Model 3: OR=0.60, 95% CI=0.40-0.91).

**Conclusion:** Increasing the daily intake of dodecanoic acid in the diet may reduce the likelihood of cataract development. Appropriate supplementation of dodecanoic acid in the daily diet can help prevent the occurrence and progression of cataracts.

## 1. Introduction

Cataract, defined as lens opacity, is one of the leading causes of visual impairment and blindness worldwide, significantly affecting patients’ vision and quality of life^[1]^. According to the World Health Organization (WHO), the global Disability-Adjusted Life Years (DALYs) for cataracts increased by 91.2% from 1990 to 2019^[2]^, with approximately 15 million people worldwide blinded by cataracts^[3]^. With the accelerating global population aging, the incidence rate of cataract has been increasing annually, emerging as a significant issue in global public health^[4–5]^. Existing epidemiological studies have identified multiple risk factors for cataract development, including individual differences, lifestyle, and environmental factors such as age, obesity, diabetes, smoking, alcohol consumption, and socioeconomic status^[6–7]^.

Surgery is currently the only treatment for cataracts. Although surgery can effectively improve the vision of cataract patients, it is highly likely to cause a series of postoperative complications, including corneal edema, fibrous membrane formation, and posterior capsule opacification^[8]^. Additionally, the high cost of surgery and limited medical conditions in some areas have greatly restricted the global feasibility of cataract surgery^[9]^.

Fatty acids are essential nutrients for the human body. Daily dietary fatty acids can be broadly classified into saturated fatty acids, monounsaturated fatty acids, and polyunsaturated fatty acids^[10]^. Dodecanoic acid, also known as lauric acid, is a common saturated medium-chain fatty acid (MCFA) widely found in coconut oil, palm oil, and breast milk^[11–14]^. When these foods are ingested, triglycerides are converted into dodecanoic acid in the body, which is further metabolized into dodecanoic acid glycerides, participating in the construction and maintenance of cell membrane stability^[15]^. Additionally, dodecanoic acid has rich biological activities, such as antibacterial, anti-inflammatory, and antioxidant functions^[16–18]^.

In recent years, increasing attention has been paid to the relationship between dietary factors and the occurrence and development of cataracts. As an important nutrient, the intake and type of fatty acids may be closely related to eye health. However, there are relatively few studies evaluating the relationship between fatty acid intake and cataract incidence, and the results have not been consistent^[19–22]^. To further explore the relationship between different types of fatty acids and cataract incidence, we conducted this study. Using the National Health and Nutrition Examination Survey (NHANES) database, we systematically analyzed the relationship between different types of fatty acid intakes and cataract incidence, providing scientific evidence for guiding fatty acid supplementation in the elderly and preventing cataracts.

## 2. Materials and Methods

### 2.1 Study Population

This study analyzed data from 20,497 participants from two consecutive NHANES cycles (2005–2006 and 2007–2008), which collected extensive information on health status, lifestyle, and nutritional intake of people from different regions, races, and age groups in the United States using a complex multi-stage probability sampling method. The survey procedures and protocols were approved by the NCHS Research Ethics Review Committee, and all participants provided informed consent^[23]^.During the sample selection process, participants under 60 years of age (n=16,773), those lacking complete cataract information (n=4), complete dietary information (n=701), and complete information on other covariates (n=483) were excluded. Finally, 2,536 participants were included in the study population(Fig.1).

**Fig. 1.**
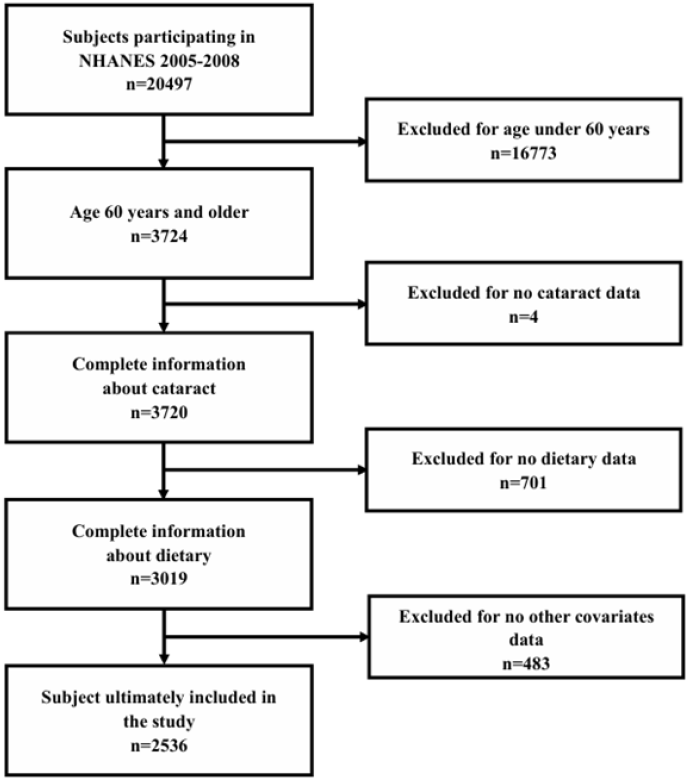
The screening process of the included studies. Flow chart of procedures from identification of eligible patients to final inclusion.

### 2.2 Cataract Identification Criteria

The diagnosis of cataract in this study was based on a direct questioning method. Participants were asked the question: “Ever had a cataract operation?” (corresponding to questionnaire question number VIQ071, with answer options “Yes” or “No”). Participants who answered “Yes” were considered to have a cataract. Data from participants who gave ambiguous answers or did not respond were excluded from the analysis. Given the increasing trend of cataract surgery rates in the United States in recent years and the more relaxed surgical criteria, this method of defining cataracts through self-reported cataract surgery is reasonable and reliable and has been used in other studies^[24–27]^.

### 2.3 Determination of Fatty Acid Intake

To more accurately reflect participants’ daily eating habits, each participant in this study participated in two 24-hour dietary recall interviews. The first dietary recall interview was conducted face-to-face at the Mobile Examination Center (MEC), and the second interview was conducted via telephone 3 to 10 days later. Daily nutrient intakes were calculated using the United States Department of Agriculture’s Food and Nutrient Database for Dietary Studies. The average of these two daily records was used to represent each participant’s dietary intake. This study included all available dietary fatty acid data from NHANES, which helps in further exploring the potential relationship between eating habits and health status.

### 2.4 Covariates Assessment

General demographic characteristics, including age, gender, race, household income, and marital status, were obtained through household interviews. Participants’ smoking and drinking statuses were also collected. Household income was divided into “≤130%”, “130%-350%”, and “>350%” based on the family income-to-poverty ratio (FPL). Smoking status was classified as “Never smoked”, “Former smoker”, and “Current smoker” based on whether the participant had smoked more than 100 cigarettes in their lifetime and whether they were still smoking at the time of the survey [28]. Body mass index (BMI) was calculated by dividing weight (kg) by the square of height (m^2^) based on physical examination results. C-reactive protein (CRP) and serum total cholesterol data were obtained from laboratory tests. Self-reported health status was obtained through a combination of physician diagnosis, relevant medication declarations, laboratory test data, and household interviews. A history of diabetes was defined as fasting blood glucose>126 mg/dL, 2-hour blood glucose in the oral glucose tolerance test (OGTT)>200 mg/dL, glycated hemoglobin (HbA1c)>6.5%, or a positive response to a diabetes history in the questionnaire. A history of cardiovascular disease (CVD) was defined as self-reported congestive heart failure, coronary heart disease, angina pectoris, or heart attack. A history of stroke (CAV) was defined as self-reported stroke. A history of hypertension (HBP) was defined as systolic blood pressure≥140 mmHg and/or diastolic blood pressure≥90 mmHg, self-reported hypertension history, or current use of antihypertensive medication. Hypercholesterolemia was defined as self-reported hypercholesterolemia history or current use of anti-hypercholesterolemia medication.

### 2.5 Statistical Analysis

All statistical analyses were performed using R 4.4.1. Given the detailed multi-stage probability sampling method used in NHANES, appropriate weights were applied to all analyses to account for the complexity of the survey design. In descriptive statistics, continuous variables were expressed as mean and standard error (SE), and categorical variables were expressed as weighted percentages. Student’s t-test and pearson chi-square test were used to compare statistical differences between the surveyed populations. Weighted multivariate logistic regression analysis was used to determine the association between various fatty acid intakes and cataract incidence. To enhance the robustness of the study results, the results were adjusted for multiple covariates, and the adjusted odds ratios (OR) and 95% confidence intervals (CI) were presented. Model 1 was adjusted for age, gender, race, household income, and marital status. Model 2 was further adjusted for BMI, smoking status, and drinking status based on Model 1. Model 3 was further adjusted for diabetes history, stroke history, and cardiovascular disease history based on Model 2. Due to the significant association observed between dodecanoic acid intake and cataract incidence, quartile regression analysis was further performed to divide the intake levels of dodecanoic acid into quartiles and study the association between dodecanoic acid intake and cataract incidence. All statistical tests were two-tailed, and a P-value of 0.05 or smaller was considered significant.

## 3. Results

### 3.1 Description of Baseline Information of the Study Sample

Tab.1 shows the baseline data and other characteristic data of participants with and without cataracts. A total of 2,536 elderly individuals aged 60 years and older were included, including 658 participants with cataracts and 1,878 participants without cataracts. After weighting, the number of cataract participants accounted for 24.85% of the total participant population. Compared with non-cataract participants, cataract participants were older, had a higher proportion of females, lower household income, lower proportion of married or cohabiting individuals, higher proportion of non-Hispanic whites, higher proportion of smokers and drinkers, higher BMI, and higher proportions of having a history of diabetes, stroke, and cardiovascular disease. CRP, serum total cholesterol, hypertension, and hypercholesterolemia had no significant impact on cataracts.

**Tab. 1.** Baseline information for the study sample.

| Characteristics | Total (n=2536) | non-Cataract (n=1878) | Cataract (n=658) | *P*-value |
| --- | --- | --- | --- | --- |
| Weighted sample size (N) | 43863222 | 32961185 | 10902037 |  |
| Continuous variables, mean (SE) |  |  |  |  |
| Age (year) | 70.01 (0.19) | 68.18 (0.19) | 75.54 (0.37) | <0.001 |
| Serum cholesterol (mg/dL) | 199.37 (1.19) | 200.63 (1.38) | 195.56 (2.25) | 0.058 |
| CRP (mg/dL) | 0.47 (0.021) | 0.46 (0.031) | 0.49 (0.037) | 0.649 |
| Category variables, n(weighted%) |  |  |  |  |
| Gender |  |  |  | <0.001 |
| Male | 1280 (44.11) | 981 (46.67) | 299 (36.36) |  |
| Female | 1256 (55.89) | 897 (53.33) | 359 (63.64) |  |
| Household income |  |  |  | <0.001 |
| ≤130% | 652 (16.60) | 481 (15.70) | 171 (19.32) |  |
| 130%-350% | 1146 (46.93) | 799 (43.47) | 347 (57.38) |  |
| >350%FPL | 738 (36.47) | 598 (40.83) | 140 (23.30) |  |
| Marital status |  |  |  | <0.001 |
| Unmarried or other | 1001 (37.43) | 696 (34.52) | 305 (46.23) |  |
| Married or living with partner | 1535 (62.57) | 1182 (65.48) | 353 (53.77) |  |
| Race |  |  |  | 0.003 |
| Mexican American | 316 (4.19) | 271 (4.66) | 45 (2.77) |  |
| Non-Hispanic White | 1545 (83.13) | 1060 (81.54) | 485 (87.95) |  |
| Non-Hispanic Black | 479 (8.04) | 393 (8.79) | 86 (5.77) |  |
| Other | 196 (4.64) | 154 (5.01) | 42 (3.51) |  |
| Smoking status |  |  |  | 0.023 |
| Never | 1149 (44.37) | 851 (44.36) | 298 (44.40) |  |
| Former | 1065 (42.65) | 758 (41.17) | 307 (47.14) |  |
| Current | 322 (12.98) | 269 (14.47) | 53 (8.46) |  |
| Drinking status |  |  |  | 0.026 |
| Never | 460 (15.50) | 321 (14.26) | 139 (19.24) |  |
| Former | 455 (18.28) | 328 (17.38) | 127 (21.02) |  |
| Current | 1621 (66.22) | 1229 (68.36) | 392 (59.74) |  |
| BMI (Kg/m<sup>2</sup>) |  |  |  | 0.002 |
| <24.0 | 485 (19.43) | 334 (18.25) | 151 (23.02) |  |
| 24.0-27.9 | 749 (30.65) | 549 (29.72) | 200 (33.44) |  |
| ≥28.0 | 1302 (49.92) | 995 (52.03) | 307 (43.54) |  |
| DM |  |  |  | 0.001 |
| No | 1773 (74.89) | 1347 (77.03) | 426 (68.44) |  |
| Yes | 763 (25.11) | 531 (22.97) | 232 (31.56) |  |
| CVA |  |  |  | <0.001 |
| No | 2324 (81.81) | 1753 (84.21) | 571 (74.56) |  |
| Yes | 212 (18.19) | 125 (15.79) | 87 (25.44) |  |
| CVD |  |  |  | <0.001 |
| No | 2039 (92.13) | 1554 (94.27) | 485 (85.67) |  |
| Yes | 497 (7.87) | 324 (5.73) | 173 (14.33) |  |
| HBP |  |  |  | 0.058 |
| No | 667 (27.51) | 518 (28.80) | 149 (23.59) |  |
| Yes | 1869 (72.49) | 1360 (71.20) | 509 (76.41) |  |
| Hypercholesterolemia |  |  |  | 0.718 |
| No | 1267 (48.09) | 945 (47.79) | 322 (49.00) |  |
| Yes | 1269 (51.91) | 933 (52.21) | 336 (51.00) |  |

### 3.2 Description of Daily Dietary Fatty Acid Intake of the Study Sample

Tab.2 shows the comparison of fatty acid intakes between cataract participants and non-cataract participants. Non-cataract participants had significantly higher total fatty acid intake than cataract participants (69.80 g vs. 62.46 g, p = 0.005). Additionally, non-cataract participants had significantly higher daily intakes of total saturated fatty acids (23.06 g vs. 20.60 g, p = 0.002), total monounsaturated fatty acids (25.53 g vs. 22.76 g, p = 0.009), and total polyunsaturated fatty acids (15.16 g vs. 13.61 g, p = 0.014) than non-cataract participants. Most saturated fatty acids, monounsaturated fatty acids, and polyunsaturated fatty acids also showed the same trend.

**Tab. 2.**
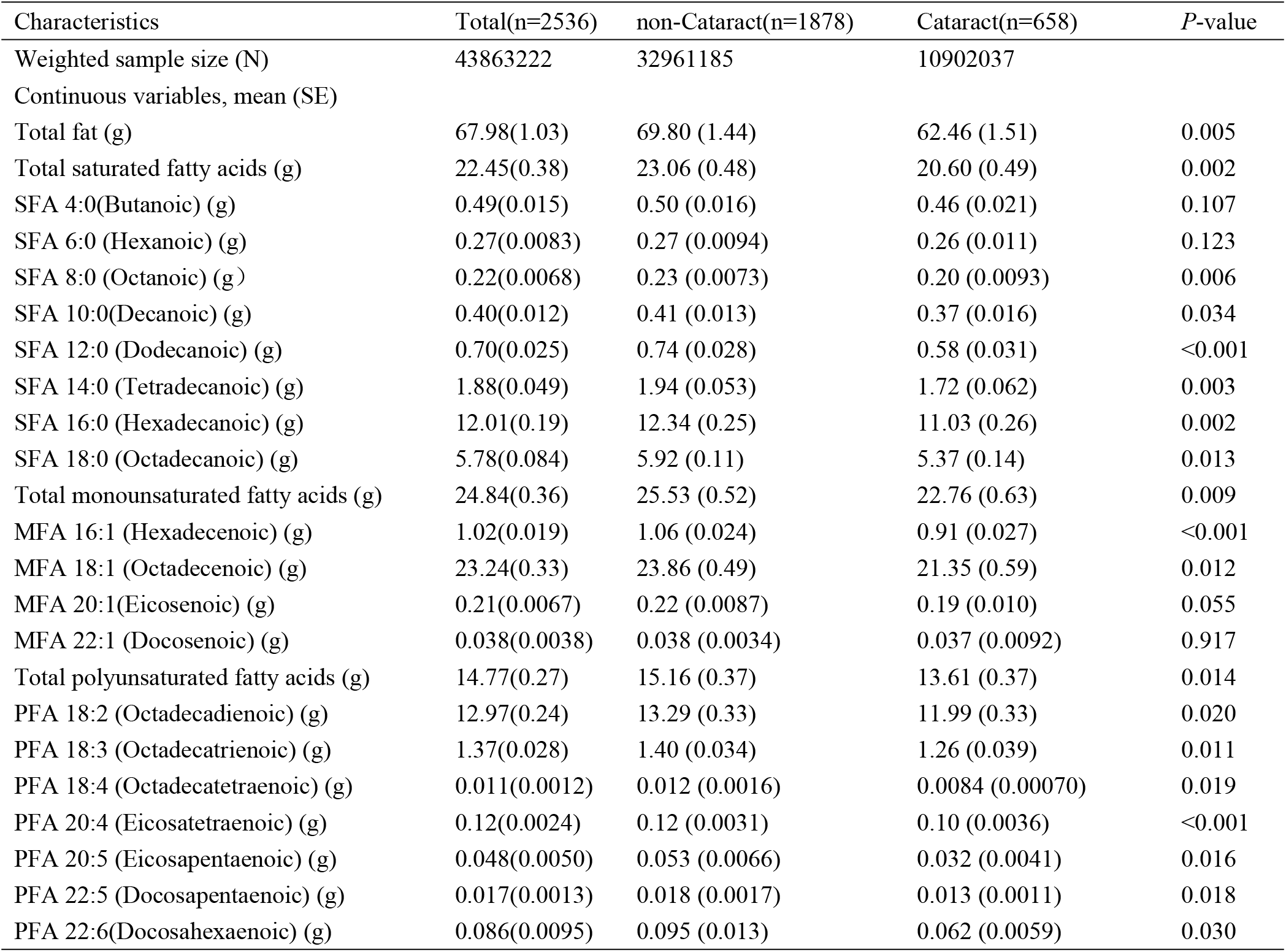
Daily dietary fatty acid intake of participants with and without cataract.

### 3.3 Association between Daily Dietary Fatty Acid Intake and the Presence of Cataract

Tab.3 shows the association between daily dietary fatty acid intake and cataract using the weighted multivariate logistic regression model. In all models, dodecanoic acid intake showed a significant negative correlation with cataract incidence (Model 1: OR = 0.79, 95% CI = 0.65–0.97; Model 2: OR = 0.80, 95% CI = 0.65–0.97; Model 3: OR = 0.81, 95% CI = 0.66–0.99). The intakes of other fatty acids were not significantly correlated with cataract incidence.

**Tab. 3.** Association between intake of daily dietary fatty acid intake and cataract.

| Characteristics | Model1<sup>a</sup> |  | Model2<sup>b</sup> |  | Model3<sup>c</sup> |  |
| --- | --- | --- | --- | --- | --- | --- |
|  | OR(95%CI) | *P*-value | OR(95%CI) | *P*-value | OR(95%CI) | *P*-value |
| SFA 8:0 (Octanoic) (g) | 0.54(0.25,1.18) | 0.115 | 0.54(0.25,1.17) | 0.111 | 0.53(0.25,1.14) | 0.094 |
| SFA 10:0(Decanoic) (g) | 0.70(0.43,1.14) | 0.145 | 0.70(0.43,1.14) | 0.143 | 0.70(0.42,1.14) | 0.138 |
| SFA 12:0 (Dodecanoic) (g) | 0.79(0.65,0.97) | 0.024 | 0.80(0.65,0.97) | 0.028 | 0.81(0.66,0.99) | 0.048 |
| SFA 14:0 (Tetradecanoic) (g) | 0.91(0.80,1.04) | 0.146 | 0.91(0.80,1.04) | 0.144 | 0.91(0.80,1.03) | 0.132 |
| SFA 16:0 (Hexadecanoic) (g) | 0.98(0.94,1.02) | 0.308 | 0.98(0.94,1.02) | 0.309 | 0.98(0.94,1.02) | 0.297 |
| SFA 18:0 (Octadecanoic) (g) | 0.97(0.90,1.05) | 0.435 | 0.97(0.89,1.06) | 0.452 | 0.97(0.89,1.06) | 0.464 |
| MFA 16:1 (Hexadecenoic) (g) | 0.83(0.58,1.18) | 0.280 | 0.82(0.57,1.20) | 0.293 | 0.81(0.55,1.19) | 0.247 |
| MFA 18:1 (Octadecenoic) (g) | 0.99(0.97,1.02) | 0.590 | 0.99(0.97,1.02) | 0.569 | 0.99(0.97,1.02) | 0.548 |
| PFA 18:2 (Octadecadienoic) (g) | 0.99(0.97,1.02) | 0.659 | 0.99(0.96,1.02) | 0.580 | 0.99(0.96,1.02) | 0.573 |
| PFA 18:3 (Octadecatrienoic) (g) | 0.88(0.75,1.02) | 0.088 | 0.86(0.73,1.02) | 0.076 | 0.87(0.73,1.03) | 0.105 |
| PFA 18:4 (Octadecatetraenoic) (g) | 0.052(0.00049,5.48) | 0.201 | 0.045(0.00047,4.43) | 0.172 | 0.056(0.00038,8.21) | 0.232 |
| PFA 20:4 (Eicosatetraenoic) (g) | 0.42(0.055,3.23) | 0.388 | 0.45(0.052,3.89) | 0.443 | 0.40(0.045,3.57) | 0.379 |
| PFA 20:5 (Eicosapentaenoic) (g) | 0.36(0.075,1.76) | 0.197 | 0.35(0.065,1.85) | 0.198 | 0.38(0.068,2.13) | 0.245 |
| PFA 22:5 (Docosapentaenoic) (g) | 0.062 (0.00014,20.76) | 0.355 | 0.054(0.000072,1.85) | 0.365 | 0.076(0.000082,70.23) | 0.427 |
| PFA 22:6(Docosahexaenoic) (g) | 0.56(0.18,1.78) | 0.312 | 0.54(0.15,1.87) | 0.305 | 0.58(0.16,2.10) | 0.376 |

Add: aModel 1: adjusted for age, race, gender, household income, marital status. bModel 2: further adjusted for BMI, smoking status, drinking status. cModel 3: further adjusted for DM, CVA, CVD.

### 3.4 Relationship of Different Quartiles of Dodecanoic Acid with the Presence of Cataract

Tab.4 shows the association analysis between different quartiles of dodecanoic acid intake and cataract. In all models, the fourth quartile of dodecanoic acid intake was significantly negatively correlated with cataract incidence (Model 1: OR = 0.58, 95% CI = 0.39-0.86; Model 2: OR = 0.58, 95% CI = 0.39-0.86; Model 3: OR = 0.60, 95% CI = 0.40-0.91).

**Tab. 4.**
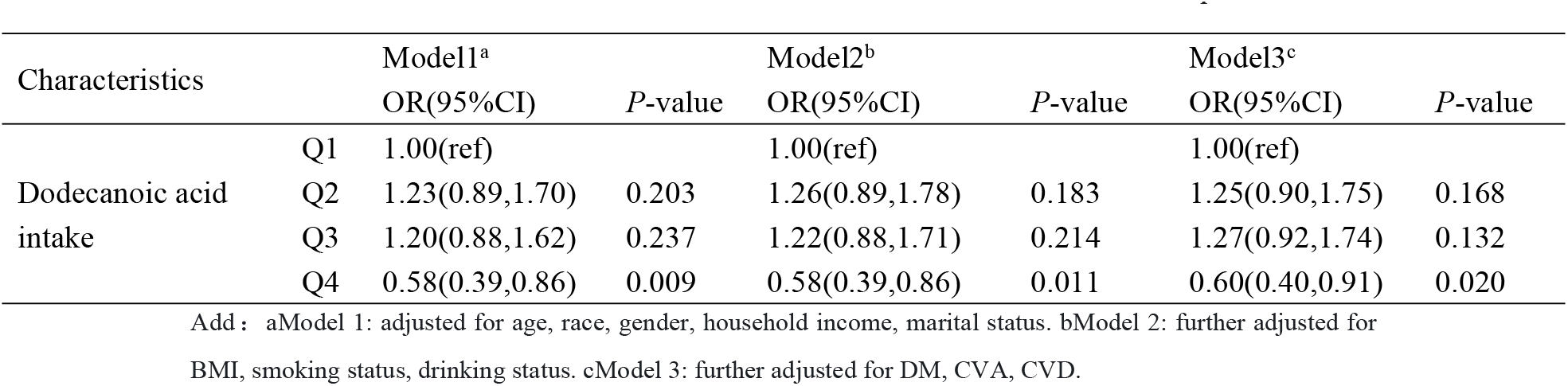
Association between dodecanoic intake levels and cataract in different quartiles.

## 4. Discussion

In this cross-sectional study, we examined the association between different fatty acid intakes and cataract incidence by extracting data from 2,536 participants in the NHANES database. The results showed a significant negative correlation between dodecanoic acid intake and cataract incidence in the weighted multivariate logistic regression analysis, indicating that participants with higher daily dietary dodecanoic acid intake had a lower likelihood of developing cataracts. We further confirmed this conclusion through quartile grouping analysis.

Dodecanoic acid is naturally present in various plant and animal oils or fats, especially in coconut oil. Coconut oil contains approximately 84% saturated fatty acids (SFA), of which about 47% is dodecanoic acid with 12 carbon atoms^[29]^. After dodecanoic acid is ingested, it is directly transported to the liver via the portal vein. Instead of being stored in adipose tissue, it is metabolized into energy and other byproducts such as ketones, which are direct energy sources for extrahepatic tissues such as the brain, heart, and muscles^[30]^.

Increasing studies have shown that dodecanoic acid not only serves as an important component of cell membranes, helping to maintain membrane fluidity and structural integrity, but also exerts multi-dimensional biological effects in the biological field. Its roles involve key biological processes such as epigenetic regulation, metabolic reprogramming, anti-inflammatory and anti-cancer effects, and drug delivery^[31]^. Studies have shown that dodecanoic acid can induce histone H3K4 trimethylation (H3K4me3) in neuroblastoma (SH-SY5Y), upregulate the expression of long non-coding RNA HOTAIR, activate the NF-κB/GLUT1 pathway to promote glucose uptake, and significantly induce apoptosis of tumor cells (BCL-2 inhibition rate of 3.6 times) through ROS-mediated mitochondrial dysfunction and BAX/Caspase-3 pathway activation^[32]^. Dodecanoic acid can inhibit hypoxia-inducible factor HIF-1α and the glycolytic key enzyme LDHA, reversing gemcitabine resistance, highlighting its dual regulatory potential in metabolism and apoptosis^[33]^. Additionally, dietary supplementation with 1% dodecanoic acid can enhance fatty acid oxidation (CPT1a upregulation) and gluconeogenesis capacity (PEPCK activity increased by 2 times) in mouse livers, enhance lipolysis in adipose tissue (ATGL activity increased by 30%), increase muscle mitochondrial DNA copy number by 40%, extend aerobic endurance by 50%, and accelerate lactate clearance by 20% by activating the AMPK-ACC pathway^[34]^. In the regulation of inflammation and tumor microenvironment, dodecanoic acid can block the TLR4/MyD88/NF-κB pathway, significantly reducing TNF-α and IL-6 levels (by 2.1 times) in LPS-induced liver injury models^[35]^. In neuroprotection and reproductive repair, dodecanoic acid reduces oxidative damage (SOD activity increased by 50%) in hyperglycemic stroke models by activating the Nrf2/ARE pathway^[36]^, and can reverse testicular oxidative stress (MDA reduced by 50%) in diabetic rats to repair sperm quality^[37]^. Based on the rich potential biological activity value of dodecanoic acid and the fact that many studies are currently exploring its preventive and therapeutic effects in various diseases, dodecanoic acid shows broad prospects from basic research to clinical applications.

The pathogenesis of cataracts is extremely complex, and the specific mechanisms underlying their development are still not fully understood. However, some studies suggest that oxidative stress is a core driver of cataract development. Excessive accumulation of reactive oxygen species (ROS) leads to the depletion of endogenous antioxidants such as glutathione (GSH) and vitamin C. The imbalance between oxidation and anti-oxidation systems in the body mediates oxidative stress, altering cell signaling pathways involved in cell proliferation and death, causing protein and lipid peroxidation in the lens, and ultimately leading to the characteristic protein aggregation, cross-linking, and light scattering observed in the lenses of cataract patients, thus causing cataract development^[38–39]^. Additionally, age-related lens protein modifications, such as post-translational modifications, mutations, or aggregation of α-crystallin, can directly disrupt its chaperone function, form β/γ-crystallin aggregates, and disrupt the ordered arrangement of lens fiber cells, leading to reduced transparency^[40]^. Metabolic disorders may amplify oxidative damage effects in diabetic cataracts through the activation of the polyol pathway or mitochondrial dysfunction, leading to NADPH/GSH imbalance^[41]^.

Therefore, dodecanoic acid may exert its inhibitory or delaying effects on the occurrence and progression of cataracts through multiple pathways: directly or indirectly scavenging intracellular free radicals via its antioxidant properties; enhancing the endogenous antioxidant system by upregulating the activities of antioxidant enzymes such as superoxide dismutase (SOD) and glutathione (GSH); neutralizing oxidative substances like reactive oxygen species (ROS); maintaining the balance between the oxidative and antioxidant systems; and inhibiting protein and lipid peroxidation in the lens to reduce oxidative damage. However, to date, studies on the relationship between dietary fatty acid intake and cataract incidence remain highly controversial. Therefore, the impact of dodecanoic acid intake on cataract incidence requires further exploration, and its specific molecular mechanisms also need to be further studied and confirmed.

In the quartile regression analysis of this study, low-dose dodecanoic acid intake was not significantly associated with cataracts, while high-dose intake showed a significant negative correlation. This may be related to the following factors: First, there may be a threshold-dependent characteristic between dodecanoic acid intake and cataract incidence. When dodecanoic acid is at a low-dose intake level, its biological effects are weak, and its potential biological effects may not be able to substantially regulate and improve the ocular cell microenvironment and related physiological processes, thus possibly unable to have a significant impact on the occurrence and development of cataracts. When dodecanoic acid intake is at a high level, it may deeply participate in the fine regulation of lens cell membrane fluidity and stability, prompting significant changes in related metabolic pathways and biological signal transduction pathways, thereby effectively reducing the risk of cataracts by optimizing the lens cell microenvironment, showing a clear negative correlation. Second, dodecanoic acid may have synergistic or antagonistic relationships with other dietary nutrients. There are complex interactions between various nutrients in the diet.

At low-dose dodecanoic acid intake, the content and effects of other nutrients may mask the potential impact of dodecanoic acid on cataracts. At high-dose intake, dodecanoic acid may either override the interference of other nutrients or exert synergistic effects with them, collectively influencing the occurrence and progression of cataracts and thus exhibit a significant negative correlation. Therefore, the specific mechanisms underlying the protective effects of dodecanoic acid against cataracts remain to be further explored.

This study benefited from a reasonable sampling design to obtain a large sample size and a nationally representative population, generating new and credible data that supports the association between fatty acid intake and cataract incidence at the overall study result level. However, this study still has certain limitations. First, due to the observational and cross-sectional nature of the NHANES database study, we cannot establish a causal relationship between dodecanoic acid and cataracts, which may hinder our interpretation of the study results. Prospective cohort studies and experimental studies are still needed to confirm the causal relationship. Second, dietary intake was self-reported through 24-hour dietary recall, which may be subject to recall bias and misclassification bias. Third, the diagnosis of cataracts in this study was based on self-reported cataract surgery. Although this method has been widely used as a good indicator of clinical cataracts, there is a risk of underestimating the disease burden of cataracts. Fourth, although this study adjusted for some confounding factors, the possibility of residual confounding factors cannot be excluded. Finally, this study used a research population based on the NHANES database. Given the numerous differences in race, socioeconomic status, and dietary structure among different countries, the validity of extrapolating the study results needs to be further evaluated.

## 5. Conclusion

Fatty acid intake is strongly correlated with cataract incidence, and there is a significant negative correlation between dodecanoic acid intake and cataracts, suggesting that appropriate supplementation of dodecanoic acid may help prevent the occurrence and development of cataracts.

## Data availability statement

Publicly available datasets were analyzed in this study. The data can be found in the NHANES repository: [https://www.n.cdc.gov/nchs/nhanes/continuousnhanes/default.aspx?BeginYear=2005] and [https://www.n.cdc.gov/nchs/nhanes/continuousnhanes/default.aspx?BeginYear=2007]. Additional data relevant to this study can be obtained by contacting the corresponding author at any time.

## Author contributions

Conceptualization, Luo Cheng and Qiu Bin; Data curation, Li Dou; Formal analysis, Xu kai; Methodology, Luo Cheng, Xu kai and Qiu Bin; Resources, Luo Cheng; Software, Xu kai and Yang feng; Validation, Yang feng and Li Dou; Writing – original draft, Xu kai; Writing – review & editing, Luo Cheng and Qiu Bin.

## Acknowledgments

We sincerely thank each of the authors for their contribution to this study also thank the Northern Medicine and Functional Food Characteristic Discipline Program of Heilongjiang Province for their help.

